# Effects of 12 weeks of Multi-nutrient supplementation on the Immune and Musculoskeletal systems of Older Adults in Aged-Care (The Pomerium Study): Protocol for a Randomised Controlled Trial

**DOI:** 10.1101/2022.01.23.22269669

**Authors:** Ahmed Al Saedi, Ben Kirk, Sandra Iuliano, Jesse Zanker, Sara Vogrin, Lata Jayaram, Shane Thomas, Christine Golding, Diana Navarro-Perez, Petra Marusic, Sean Leng, Ralph Nanan, Gustavo Duque

## Abstract

**Introduction:** Immunosenescence leads to increased morbidity and mortality associated with viral infections and weaker vaccine responses. This has been well documented for seasonal influenza and the current pandemic with Sars-Cov2, which disproportionately impact older adults, particularly those in residential aged care facilities. Inadequate nutrient intake associated with impaired immunity, respiratory and muscle function are likely to augment the effects of immunosenescence. In this study, we test whether the effects of inadequate nutrition can be reversed by multi-nutrient supplementation, consequently enhancing vaccine responses, reducing the risk of viral infections, and improving respiratory and muscle function.

**Methods and analysis:** The Pomerium Study is a 12-week, single-blinded, randomised, placebo-controlled trial testing the effects of two daily servings of an oral multi-nutrient supplement (330 kcal, 20g protein, 1.2g CaHMB, 449mg calcium, 520IU vitamin D_3_, and 25 vitamins and minerals) on the immune system and muscle and respiratory function of older adults in aged-care in Melbourne, Australia. 160 older adults (≥75 years old) will be recruited from aged-care facilities and randomised to treatment (multi-nutrient supplement) or control (usual care). Primary outcome is the change in T-cell subsets CD8+ and CD28null counts at 4 and 12 weeks post-intervention. Secondary outcomes measured at baseline and after 12 weeks post-intervention are multiple markers of immunosenescence, body composition (bioimpedance), handgrip strength (dynamometer), physical function (short physical performance battery), respiratory function (spirometry), and quality of life (EQ-5D-3L). Incidence and complications of COVID-19 and/or viral infections (i.e., hospitalisation, complications, or death) will be recorded throughout the trial.

**Discussion:** If the Pomerium Study demonstrates efficacy and safety of a multi-nutrient supplement on immune, muscle and respiratory function, it may be suitable as a strategy to reduce the adverse outcomes from seasonal influenza and viral infections such as COVID-19 in older adults in aged-care.

**Funding, Ethics, Registration and Dissemination:** The study is funded by the Australian Medical Research Future Fund. It is approved by Melbourne Health Human Research Ethics Committee (Ref No. HREC/73985/MH-2021, ERM Ref No. RMH73985, Melbourne Health Site Ref No. 2021.115), and registered at ANZCTR (12621000420842). Results will be published in peer-reviewed journals and made available to aged-care stakeholders, including providers, residents, and government bodies.

**Article Summary Strengths and Limitations:**

- This is the first study performing a comprehensive immune, respiratory and functional assessment in aged care residents after receiving a multi-nutrient solution that is commercially available.
- Our design and tested intervention assure that the results of the study will be rapidly translated into practice.
- The main limitation is that any biological effect observed cannot be attributed to one component of the multi-nutrient supplement.
- Another limitation is that the potential effect of group differences in energy intake on outcomes can only be monitored by assessing regular dietary intake and weight changes during the study period.

## INTRODUCTION

Ageing is characterised by a decline in immune function known as immunosenescence.^1^ This process reduces resistance to infectious diseases (e.g., pneumonia, influenza, meningitis, and urinary tract infections).^2^ During a pandemic, the concept of immunosenescence is of relevance as older adults, particularly those living in residential aged care facilities (RACFs), are at high risk of acquiring infectious diseases and experiencing more adverse outcomes.^2 3^ Both intrinsic and extrinsic factors contribute to the predisposition of institutionalised older adults to respiratory viral infections. Intrinsic risk factors include immunosenescence, malnutrition, low serum levels of vitamin D, limited mobility, poor muscular and respiratory function, and comorbidities.^4-6^ Extrinsic factors include lack of appropriate infection control procedures, and limited access to personal protective equipment.^7^ Vaccination to viral assaults is the primary preventative strategy to reduce both onset and severity of viral infection.^7^

Malnutrition is common in aged-care residents^8^ and is associated with compromised immune profile (i.e., lower levels of T-cell subsets (CD8+ CD28null), high NK:CD4+ T cell ratio, and low serum levels of interleukin (IL)-7).^1,8,9,10,11^ Dietary protein contains immunoglobins that protect against antigens,^12^ and vitamin D modulates immune function by stimulating the differentiation of regulatory T and B cells.^13^ Inadequate protein intakes and vitamin D deficiency are common in older adults in aged care.^14-16^ On average daily protein intake of 0.8 g/kg bodyweight have been observed in older adults living in Australian residential aged care;^11^ an amount considered insufficient supporting optimal immune and musculoskeletal function.^17^ Furthermore, up to 77.5% of older adults in aged-care are vitamin D deficient (serum 25(OH)D levels < 50 nmol/L),^18,19^ and inappropriate vitamin D levels are associated with reduced severity and mortality from COVID-19.^20,21^

Sarcopenia is a progressive and generalised skeletal muscle disorder characterised by decreased muscle quality, quantity and function.^22,23^ This process is particularly evident in respiratory muscles,^24^ where it impairs the ability to produce appropriate tidal volume^25^ and perform high force expulsive airway clearance manoeuvres.^26^ Multiple studies have shown that area of the pectoralis, psoas, and paravertebral muscles on cross-sectional CT images is associated with lean muscle mass, handgrip strength, sarcopenia, and health.^23,27,28^ Sarcopenia was also evident in COVID-19 patients.^29,30^ Baseline sarcopenia was independently associated with a prolonged hospital stay in patients with COVID-19.^31^ Higher paraspinal muscle radiodensity, a proxy measure of lower muscle fat deposition, was associated with a reduced risk of disease deterioration and decreased likelihood of prolonged viral shedding among female patients with severe COVID-19.^32^ In addition to the well-known independent risk factors (i.e. age, obesity, COPD, and C-reactive protein (CRP) level), low grip strength is independently associated with increased severity of COVID-19.^33^ Moreover, decreased muscle strength is an independent risk factor for COVID-19 severity in adults 50 years of age or older.^34^ Therefore, low muscle mass and strength are considered risk factors for COVID-19 severity.^35,36^

Of the nutrients purported to support immune and musculoskeletal function, whey protein contains bioactive immunoglobins (such as lactoferrin) with immunostimulatory properties,^12^ as well as essential amino acids (in particular leucine) required for muscle protein synthesis.^37 38^ Another potent stimulator of muscle protein synthesis is calcium β-hydroxy-β-methyl-butyrate (caHMB),^39^ a leucine metabolite, which appears to offer complementary benefits to leucine by simultaneously dampening muscle proteolysis.^40^ This may be important for older adults with compromised immune function and/or sarcopenia, where chronic-low grade inflammation may be driving muscle loss.^41,42^ Vitamin D also interacts with protein to support this anabolic signalling network, and sufficient vitamin D levels (> 50 nmol/L) may be required for protein to increase muscle mass as observed in animal models and older adults with sarcopenia.^43,44^

Several other vitamins and minerals, such as calcium and iron that act as cofactors in metabolism^41,45^ may help support the immune, respiratory and musculoskeletal systems and reduce the risk of adverse events in this population, such as falls, fractures and respiratory infections.^45^ An RCT involving 157 older adults (>65 years) living in long-term care supplemented with a nutrition formula that contained triacylglycerol, protein, antioxidants, selenium, zinc, and 28 vitamins and minerals for 4 weeks, demonstrated enhanced immune function as indicated by increased influenza vaccine response and lymphocyte activation, less fever, and fewer days of symptoms of upper respiratory tract infections.^46^ Similar improvements to response to influenza and pneumococcal vaccination have been observed in older adults living in the community provided similar nutritional supplements.^47,48^ Therefore, correcting nutritional inadequacy is a viable option to support immune, muscle and respiratory function in older adults living in aged-care.

The aim of this 12-week single-blind, randomised controlled study (Protocol version 2.0) is to test the effects of two daily servings of a multi-nutrient supplement (containing whey protein, leucine, CaHMB, vitamin D_3_, calcium plus 25 vitamins and minerals) on the immune system and muscle and respiratory function of older adults in aged-care. We hypothesise that provision of this multi-nutrient supplement will improve multiple immune and functional variables and reduce the number and severity of cases of respiratory viral infections. To test this hypothesis, we propose measuring T-cell subsets (CD8+ CD28null) as our primary outcome. Our secondary outcome measures include a comprehensive immune profile (cell counts and serum cytokines), body composition (bioimpedance), handgrip strength (dynamometer), physical function (short physical performance battery), respiratory function (spirometry), and quality of life (QoL) (EQ-5D-3L).

## METHODS AND ANALYSIS

### Trial design and population

This is a 12-week parallel group single-blinded (only assessors blinded) randomised controlled trial involving 160 older adults living in aged-care, randomised to either two daily doses of a multi-nutrient supplement (treatment) (Appendix 1) or usual care (control) (Figure 1). Participants will also be assessed 12 weeks after supplementation ceases. Assessments will be performed at baseline (immune, serum, respiratory and muscle-related measures, and QoL), week 4 (immune and serum only), week 12 (immune, serum & other measures) and 12 weeks following cessation of supplementation (event recording only). The trial will follow the CONSORT guidelines for reporting randomised trials.^49^

**Figure 1.**
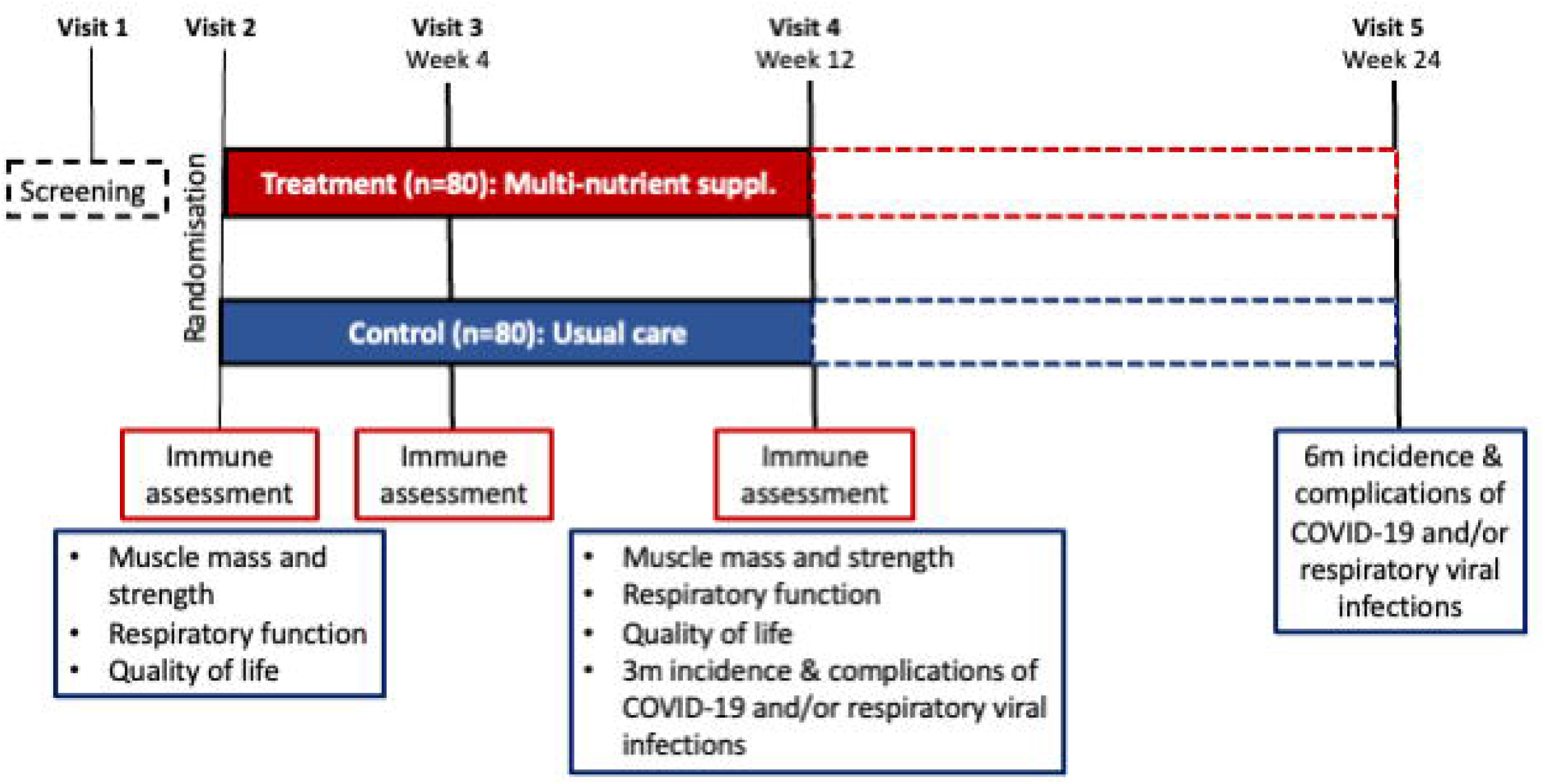
The Pomerium Study design.

### Patient and Public Involvement

The research question and methodology were based on previous experience by the investigators testing nutritional interventions in aged care residents, including surveying their preferences asking for their feedback and acceptance of the multi-nutrient solutions.^11,50^ Our Consumer Representative at the Australian Institute for Musculoskeletal Science (AIMSS) participated in the design of this protocol and is also an Associate Investigator in the grant application. Results will be disseminated via regular reports submitted to the participating aged care facilities and distributed amongst the participants and their families.

### Recruitment

RACFs managers will be contacted to determine their willingness to be involved in the trial and encourage residents’ participation. Agreement with the aged-care manager will be formalised via an agreement document between the RACF and the research institute conducting the trial. Recruitment of participants will involve promoting the trial in facility newsletters and presenting the trial to residents and their families at resident and relative meetings. This recruitment method has been successfully used in the past.^50,51^

### Trial population and randomisation

The target population is sarcopenic aged-care residents aged 75 years and older that may or may not have received a COVID-19 or seasonal influenza vaccination. Sarcopenia will be diagnosed using the Sarcopenia Definition and Outcomes Consortium criteria.^52^ In addition to the presence of sarcopenia, participants must meet the inclusion and exclusion criteria to participate (Table 1), which include capacity to participate and ingest the multi-nutrient treatment. The trial statistician will generate the randomisation sequence using permuted block design stratified by aged care facility and uploaded to research electronic data capture (REDCap). Treatment allocation will be concealed until the time of the randomisation.

**Table 1:**
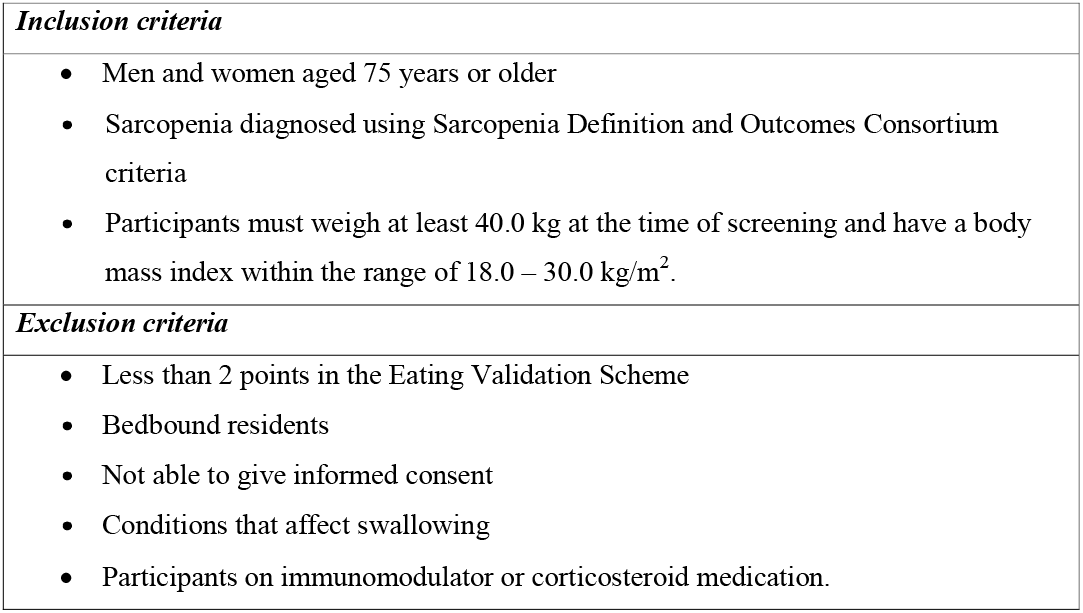
Inclusion criteria and Exclusion criteria.

|  |
| --- |
| ***Inclusion criteria*** |
| • Men and women aged 75 years or older<br>• Sarcopenia diagnosed using Sarcopenia Definition and Outcomes Consortium criteria<br>• Participants must weigh at least 40.0 kg at the time of screening and have a body mass index within the range of 18.0 – 30.0 kg/m<sup>2</sup>. |
| ***Exclusion criteria*** |
| • Less than 2 points in the Eating Validation Scheme<br>• Bedbound residents<br>• Not able to give informed consent<br>• Conditions that affect swallowing<br>• Participants on immunomodulator or corticosteroid medication. |

### Testing procedures

The visit schedule is illustrated in Figure 1, and assessments are presented in Table 2. The visit schedule consists of screening (visit 1), baseline/randomisation (visit 2), immune function (visit 3), final (visit 4), and 12-week follow up (visit 5) assessments. It is anticipated that the time commitment for participants will be between 30 to 60 minutes per visit.

**Table 2.**
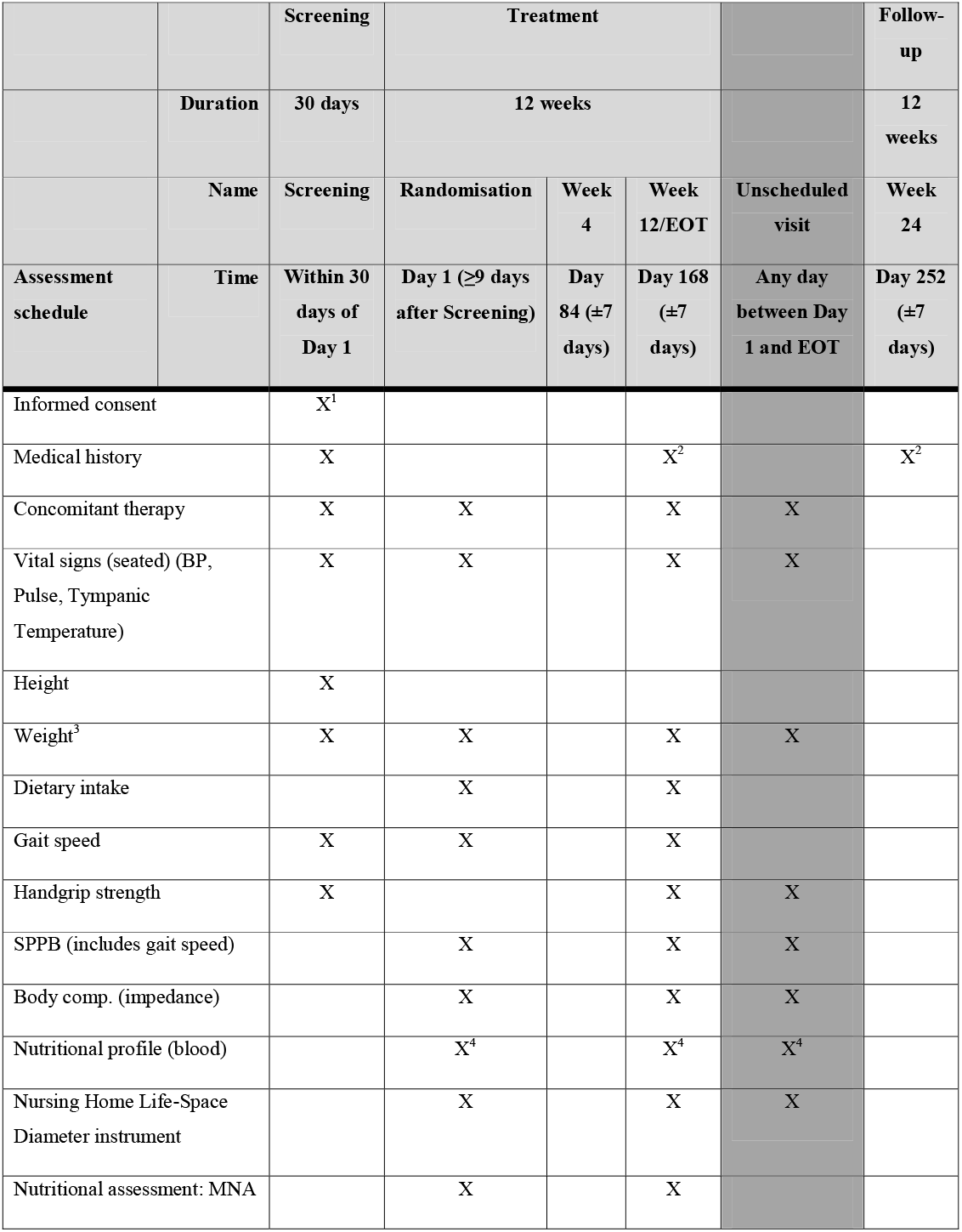
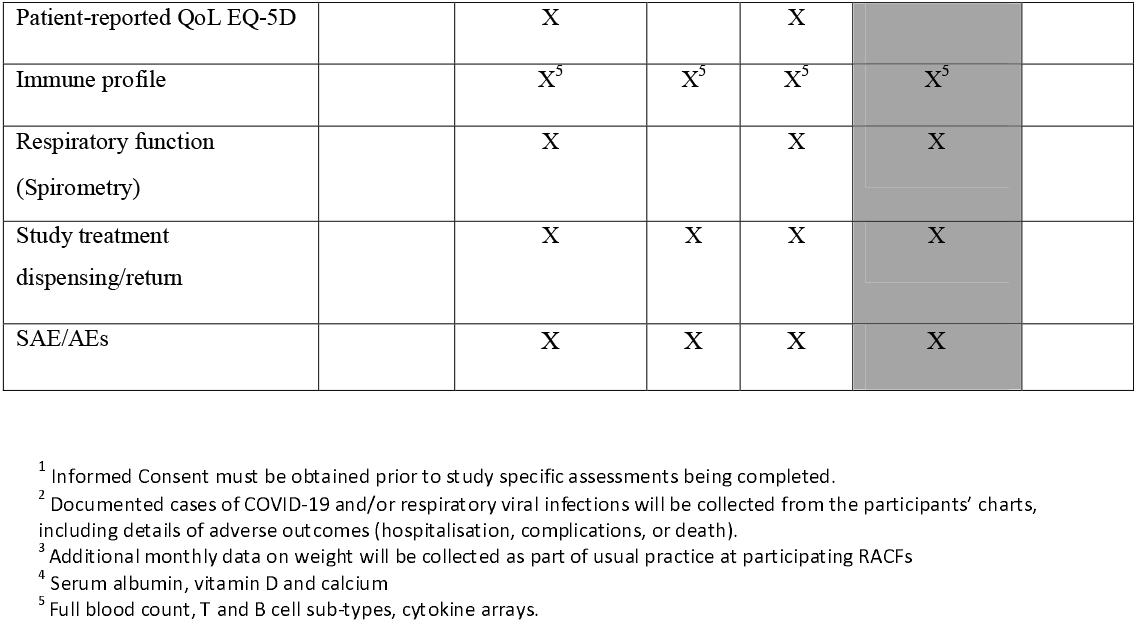
Assessment and Visit Schedule.

#### Immune and nutritional profile

Non-fasting blood (25 mL) will be collected by venepuncture by trained personnel. Twenty mL will be analysed by flow cytometry (Aurora^®^) to quantify full blood counts, T and B cell counts and their subsets (particularly T-cell subsets CD8+ and CD28null) and other surface phenotypes of immunosenescence (Table 3). Serum concentrations of 40 interleukins (Appendix 2) will be quantified at the Australian Institute for Musculoskeletal Science (AIMSS) using a MILLIPLEX MAP Human Cytokine/Chemokine Magnetic Bead Panel (Millipore). Serum vitamin D, calcium and albumin will be assayed at Dorevitch Pathology (Melbourne, Australia) using validated techniques.^53^

**Table 3.**
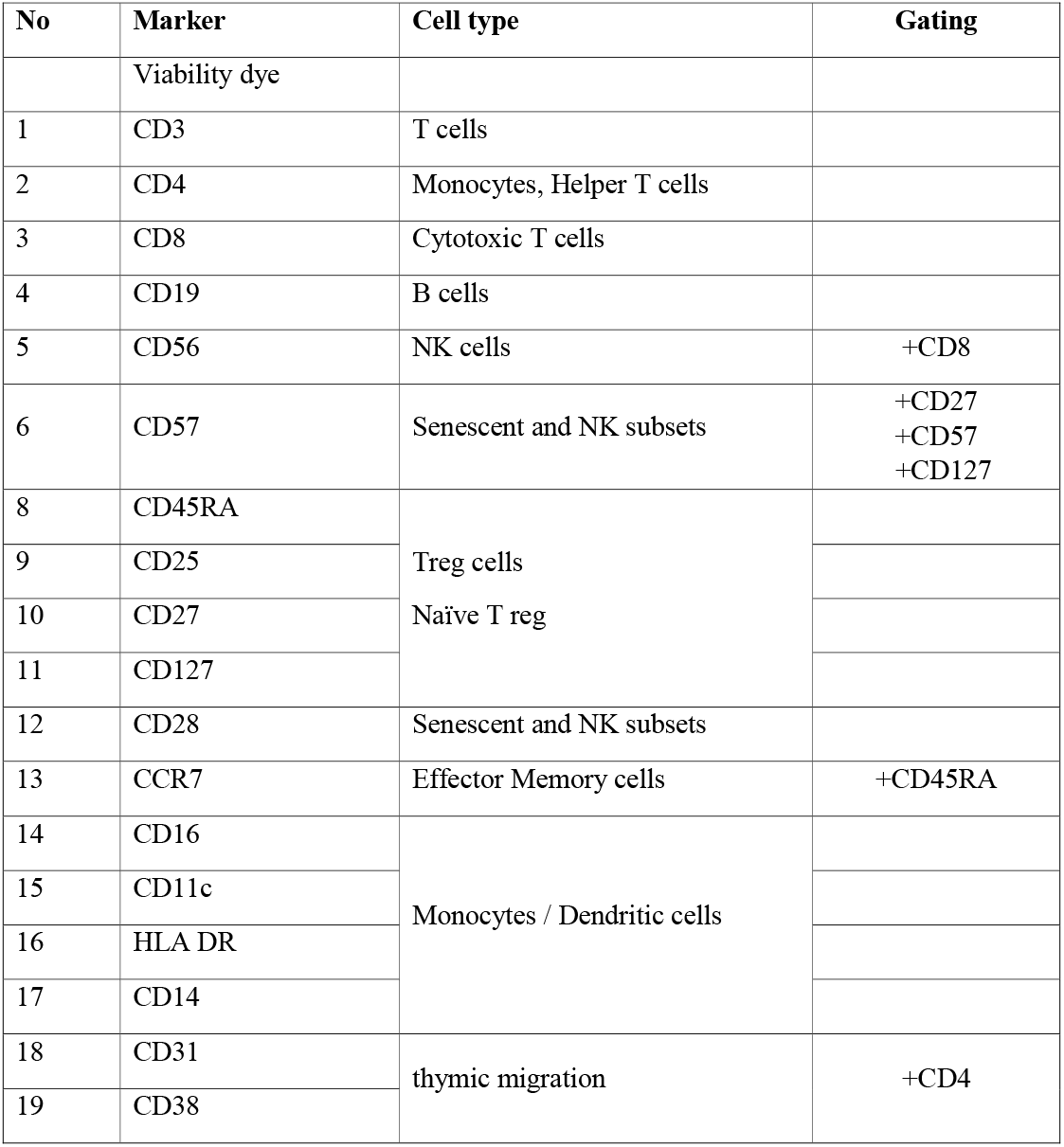
Markers of immunosenescence included in the flow cytometry analyses.

#### Physical function

##### Handgrip strength

Assessed using a Jamar hydraulic dynamometer (Sammons Preston Inc. Bolingbrook, IL). Participants will be seated with their arm resting (at 90 degrees) on the chair arm and instructed to squeeze the dynamometer at maximal effort (test is performed 3 times on each side with 30□seconds rest between each test). The highest results of three attempts will be recorded.

##### Gait Speed

Measured as the time to walk 6 meters at normal speed using a stopwatch. The use of walking aids (e.g., cane, walker) will be recorded. Three tests (3 min between) are performed with the best speed recorded.^54^

##### Short Physical Performance Battery (SPPB)

Will be used to assess lower extremity function using tasks that mimic daily activities. The SPPB examines static balance, gait speed, and lower body strength. Balance assessments are composed of three tasks that become progressively more challenging, i.e. standing unaided for 10 seconds with feet together, feet in semi tandem (one foot in front of the other foot, with the big toe of the back foot in the groove of the front foot) and full tandem position.^55^ The 5 times sit-to-stand test is performed with the participant starting in the seated position. After confirming the ability to perform one sit-to-stand action (chair arms for help with balance), participants are instructed to stand and sit 5 times as quickly as possible, ensuring feet are flat on the floor. Scores are allocated according to performance, with an overall maximum score of 12 (0-6 is low performance, 7-9 moderate and 10-12 high performance).^56^

##### Body Composition

Bioelectrical Impedance (TIDC360S, Tanita, Wedderburn, Australia) will be used to calculate body composition after calibrating the device for the following variables: weight, height, age, and sex.

##### Vital signs

Blood pressure (BP) will be measured by trained personal in a seated position using an automated electronic BP monitor. Heart rate and temperature will also be recorded.

##### Respiratory function

Spirometry (Forced Expiratory Vital Capacity in 1 second (FEV1), Forced vital capacity (FVC), and FEV1/FVC ratio) pre- and post-salbutamol will be assessed using a calibrated portable hand-held Micro spirometer (CareFusion, Kent, United Kingdom) and performed to American Thoracic Society (ATS) standards with predicted values from NHANES III. ^57^

##### Anthropometry

Height (digital stadiometer, (SECA20)) and weight (homologated electronic balance (SECA20)) will be measured with the participant barefoot (or wearing socks or stockings) and wearing light clothing. BMI (weight in kilograms divided by height in meters squared) will be calculated. Nutritional status will be assessed using the long Mini-Nutritional Assessment (MNA) tool,^58^ a validated instrument that contains 18 items and evaluates four different aspects of nutritional status: anthropometric assessment (body mass index (BMI), weight loss, and arm and calf circumferences); general assessment (lifestyle, medication, mobility and presence of signs of depression or dementia); short dietary assessment (number of meals, food and fluid intake and autonomy of feeding); and subjective assessment (self-perception of health and nutrition).

##### Quality of life (QoL)

Assessed using the EQ-5D-3L questionnaire.^59^ Briefly, the questionnaire enables comparisons of quality of life across different diseases or health states using a single score and has five dimensions (mobility, self-care, usual activities, pain/discomfort, and anxiety/depression)

##### Nursing Home Life-Space Diameter instrument

Used to identify social isolation and self-restricted life-space mobility.^60^ This instrument, which is measured over two weeks, separates a patient’s living area into four spaces: their room, outside the room but within the unit, outside the unit but within the facility, and outside the facility.

##### Dietary intake

Determined on two random days using the validated method of visual estimation of plate waste.^61^ Standard serves will be weighed on a digital food scale (±1g) (Soehnle Page Profi), and foods and beverages served and wasted will be compared against the standard serve using a seven-point scale. The 7-point scale represents portions of each food consumed (or remaining): 0=no food remaining, +M=1 mouthful remaining, 1/4=25% remaining, 1/2=50% remaining, 3/4=75% remaining, −M=1 mouthful consumed (90% remaining), 1=no food eaten. Meals served will be rated against the standard meal (medium given the value of 100%); small serving = 75%, large serving = 125%, extra-large serving = 150%. Consumption will be calculated as the difference between amounts served and wasted.

##### Medical record review

Medical history, documented cases of COVID-19 and/or respiratory viral infections will be collected from the participant’s medical records, including details of serious adverse outcomes (hospitalisation, complications, or death).

##### Adverse events (AE)

All compulsory incident reports documented at each facility will be reviewed, and any adverse events reported for study participants will be recorded. AE are defined as any untoward medical occurrence(s) in a study participant that may or may not be temporally or causally associated with the use of the multi-nutrient supplement and is considered serious if it results in death, is life-threatening, requires inpatient hospitalisation or prolongation of existing hospitalisation, or results in persistent or significant disability.

### Study Visits (Figure 1)

#### Visit 1 – Screening

- Informed Consent is obtained before study assessments commence
- Medication use (existing therapies or therapies changed or ceased in the last 3 months) will be documented.

#### Visit 2 – Randomisation

- Vital signs, weight, MNA; Physical Performance Assessment
- SPPB, gait speed and handgrip strength
- Body composition
- Nursing Home Life-Space Diameter instrument
- QoL
- Respiratory function
- Non-fasting blood sample (immune and nutritional profile)

#### Visit 3 (4 weeks)

- Non-fasting blood sample (immune and nutritional profile)
- Concomitant medication and AE record

#### Visit 4 (12 weeks) (similar assessment to visit 2)

- Document cases, severity, and outcomes of COVID-19 and/or respiratory viral infections
- Non-fasting blood sample (immune and nutritional profile)
- Concomitant medication and AE record

#### Visit 5 (24 weeks; 12 weeks after cessation of supplementation)

- Document cases, severity, and outcomes of COVID-19 and/or respiratory viral infections

### Unscheduled Visits

If deemed by the Investigators that additional assessments are required for medical or safety reasons.

### Treatment

#### Multi-nutrient manipulation and storage

After screening, eligible participants will be randomised (as described earlier) to receive two daily 220 ml bottles of the multi-nutrient supplement (intervention) (Appendix 1) or usual care (control). The multi-nutrient supplement will be supplied to the facility by an assigned investigator, handled and stored safely and properly, and kept in a secure location that only the assigned investigator and designated staff at the facility have access to. Upon receipt, the multi-nutrient supplement will be stored according to the instructions specified by the manufacturer. Where possible, the multi-nutrient supplement will be refrigerated at 5 degrees at the aged care facility prior to administration. Documentation of the dispensing process will be maintained.

### Potential side effects and monitoring

The multi-nutrient supplement is listed in the Australian Register of Therapeutic Goods. The dose used in this study is the amount recommended by the manufacturer (Abbott Pharmaceuticals, Australia). The potential risks from consuming the multi-nutrient supplement are considered similar to other commercially available supplements.

Two daily doses of the multi-nutrient supplement provide 1,000 IU of vitamin D. The Institute of Medicine has set the dose of 4000 IU per day as the Tolerable Upper Limit, so total vitamin D intake, including that from other supplements, will be recorded. Participants will be monitored for signs of toxicity (nausea, vomiting, diarrhoea, or frequent urination) and blood 25(OH)D and calcium levels evaluated at baseline and visit 4. A clinical trials monitoring committee is established at the research institute. Any AEs will be communicated to this committee, and action will be taken based on the severity of the event.

### Treatment Compliance

Compliance will be monitored weekly with all bottles returned to the research institute and unused product measured and recorded.

### Treatment blinding

Staff involved in assessments will be blinded to treatment allocation. Randomisation data will be kept confidential and only accessible by designated authorised, un-blinded study personnel. Unblinding will only occur in the case of participant emergencies and at the conclusion of the study and statistical analyses.

### Participant withdrawal

Participation will be discontinued if the investigator or the monitoring committee deems that continuing study treatment would be detrimental to a person’s well-being. This may include but is not limited to; (i) emergence of one or more adverse events or laboratory abnormalities that, in the judgment of the Investigator, prevents the person from safely continuing in the study, (ii) intentional loss of adherence for seven consecutive days (including hospitalisation); or (iii) a protocol deviation that results in a significant risk to the person’s safety. Discontinuation of participation may also occur if (i) death; (ii) discharge from the aged care facility; or (iii) withdrawal of consent by the participant.

Withdrawal of consent may be made at any time and for any reason, with details documented. Participants may withdraw consent to (i) no longer participate in the entire study, (ii) not participate in a particular part of the study or aspects of assessments, (iii) not participate in further visits or assessments, or (iv) have any further study related contact. In this case, contact would only be made for safety reasons. In the case of (i) and (iii) study treatment will be discontinued, and no further assessments conducted.

### Lost to follow-up

For participants whose status is unclear because they fail to undergo study visits without stating an intention to discontinue, the Investigator will show ‘due diligence’ by documenting steps taken to determine their absence, e.g., view medical records or inquiry through facility staff. A participant should not be considered lost to follow-up until their scheduled end of study visit has occurred. Participants who are discontinued from the study for any reason will not be replaced.

### Study completion and post-study treatment

Each participant will be required to complete the study in its entirety. The study will be considered complete when the last participant completes their final visit, and any repeat assessments associated with this visit have been documented and followed up appropriately by the Investigator.

### Outcomes

To determine the efficacy of two daily doses of a multi-nutrient supplement on immune, muscle and respiratory function of older adults in residential aged care, the following outcomes will be assessed:

### Primary outcome

T-cell subsets CD8+ and CD28null measured at baseline and weeks 4 and 12.

### Secondary outcomes

i. Multiple immunosenescence markers measured at baseline and weeks 4 and 12 (Table 3 and Appendix 2)
ii. Serum vitamin D (25OHD) and albumin, measured at weeks 4 and 12.
iii. SPPB, handgrip strength, body composition (fat and lean mass)
iv. Respiratory function (FEV1, FVC and FEV1/FVC ratio)
v. QoL and Nursing Home Life-Space Diameter instrument

### Statistics

#### Sample size calculation

Available data on normal levels of T-cell subset CD28null indicated the standard deviation for males and females aged 60-80-year-old is 16% and 21% for those aged 80-100 years.^60^ A 10% difference in T-cell subset CD28null is considered clinically relevant.^60^ Assuming a standard deviation of 20%, to detect a 10% difference with 80% power at the 5% significance level, 128 participants will be required. Allowing for 20% attrition, 160 participants (80 per group) are required.

### Statistical Analysis

Descriptive statistics will be used to summarise information collected for each outcome at each time-point. The primary outcome of the study is the comparison of T-cell subsets CD8+ and CD28null at 4 weeks and 24 weeks from baseline, which will be assessed using a generalised linear mixed model adjusted for baseline levels. If the model is of poor fit by visual inspection of residuals, the outcome will be transformed using natural logarithm. If the model fit is still insufficient, non-parametric tests will be used for analysis.

The analysis will be on an intention-to-treat basis with additional per-protocol analysis. The extent of missing data will be evaluated. Missing data will be handled within the generalised linear mixed model. Where non-parametric analysis will be performed, complete case analysis will be performed (if missing data is minimal), or simple imputation will be performed. Secondary outcomes will be analysed in a similar manner, using mixed-effects linear regression for continuous outcomes and mixed-effects negative binomial regression for counts. Statistical significance will be assumed at p□<□0.05. No adjustments for multiple comparisons will be made; however secondary outcomes will be interpreted in light of multiple comparisons.

### Data Collection and Storage

#### Data Management, Security and Handling

The Investigators are responsible for ensuring the accuracy, completeness, legibility, and timeliness of data reported. Designated research staff are provided with an individual log-in to enter data required by the protocol into the electronic Case Report Form (eCRF) within a secure electronic password-protected database (REDCap) hosted on a secure server by The University of Melbourne. All data will have an external originating source (either written or electronic). Paper-based source data will be stored in a locked office at AIMSS. Automatic validation syntax set within REDCap will check for data discrepancies and generating appropriate error messages will allow for data to be confirmed or corrected. Participants will be de-identified and given a unique participant number.

#### Data Sharing

No study data or information will be released to any unauthorised third party without prior written approval by The University of Melbourne. Recipients will treat the data according to the Australian Privacy Principles or similar privacy legislation. The recipients will not use or disclose the information untowardly or outside the parameters of the agreement between them and the Institution (The University of Melbourne). No individual will be identified in reports or publications; only group-level data will be presented.

Participants will only be identified by a unique participant number. The Investigator will maintain a confidential participant identification list that allows the unambiguous identification of each participant. All relevant and applicable laws and guidelines will be applied to any data that is leaving the Institution. After the study is complete, a study summary will be provided to participating facilities and individual participants.

#### Record Retention

All study documents will be retained for a minimum of 15 years after study completion and will be disposed of in a standard secure manner at the end of the archival period. Only authorised study staff will have access to the data.

## DISCUSSION

We aim to determine whether a 12-week multi-nutrient supplement improves immune, muscle and respiratory function in older adults living in aged-care who are at high risk of adverse outcomes due to seasonal influenza and other viral infections such as COVID-19. Older adults, particularly aged-care residents, are prioritised for influenza and COVID-19 vaccinations. However, low immune responses to standard vaccines because of immunosenescence may compromise vaccine effectiveness^6,62^ Therefore, vaccination efficacy may be improved by priming the immune system of older adults.

The proposed outcome from this work is that multi-nutrient supplementation will improve cellular immunity. Viral infections have several features that make them useful for studying T cell immune responses as the productive infection is localised to lung tissue, and no persisting virus can be detected.^63^ These features make influenza/COVID-19 infections a good model for studying both the effector and memory phases of T cell response. Multiple studies have tested proliferative responses to *in vitro* challenges with influenza antigens via examined total T cell proliferation, and it is known that proliferative responses to influenza vaccine are generally higher within CD4 than CD8 T cell subsets.^64-66^ These data indicate that specific nutritional supplements can enhance T cell proliferative responses to viral challenges. Noteworthy, measuring immune competency in older adults should involve several criteria because immune changes from ageing and malnutrition are similar; nutrient supplementation may improve immune status and clinical outcomes in older adults, ^67 68^ and at risk of sarcopenia.^69^ These criteria must include full blood counts, T and B cell counts and their function and subsets (particularly T-cell subsets CD8+, CD28null, Treg, and Dendritic cells), and concentrations of serum interleukins (particularly interleukin (IL) -7).^1,9,10,70^

As the population ages and the risk of potential pandemics remain, it is reasonable to prepare vulnerable older adults for viral assaults by implementing efficacious evidence-based interventions using specialised nutritional supplements.^40 71-74^ Nutritional interventions using vitamin D, protein, zinc and selenium enhanced anti-viral resistance against COVID-19 in older adults. ^75^ Among COVID-19 inpatients (n = 134), 19% of patients in intensive care units had serum 25(OH)D levels above 50 nmol/L compared to 39% of those in conventional medical wards.^76^ In a randomised placebo-controlled trial involving 38 older adults, vitamin D supplementation (100,000 IU/15 days) promoted a higher TGFβ plasma level (20.8ng/mL) in response to influenza vaccination and directed the lymphocyte polarisation toward a tolerogenic immune response.^77^ Furthermore, vitamin D has been associated with improved pulmonary function and reduced incidence of airway infections. An RCT of 86 older adults patients showed that consumption of 50,000 IU vitamin D supplementation in a daily diet could increase quality of life and pulmonary function in adult patients.^78^ Moreover, higher serum vitamin D levels (>50 nmol/L) among adults are associated with decreased odds of obstructive lung disease in the general population.^79^ In addition, vitamin D supplementation (2000 IU/day) reduced the risk for pneumonia, acute exacerbations of respiratory diseases, and lung function decline in older adults.^80^ Therefore, vitamin D supplementation may have a beneficial role against viral infections in aged-care residents.

Various studies indicate that older adults require a daily protein intake of 1.0 to 1.2g / kg body weight to support muscle health and function.^38,81^ In well-controlled and powered RCTs of sarcopenic older adults, supplementation using whey protein enhances muscle mass and function^82 83^ and prevents mobility impairment.^84 85^ Our team recently showed in a 2-year clustered-randomised nutrition intervention involving over 7,000 older adults in aged-care that increased dietary protein (from 0.9 to 1.1g/kg body weight) and calcium intakes were associated with maintenance of appendicular muscle mass.^50^ An *in vitro* study demonstrated that whey protein has a direct-acting inhibitor of SARS-CoV-2 infection and replication.^86^

CaHMB increases protein synthesis and prevents protein degradation in young and older adults.^39,87^. CaHMB supplementation contributed to the preservation of muscle mass in older adults and prevention of muscle atrophy induced by bed rest or other factors.^88^ Supplementation with CaHMB (3g), vitamin D (1000IU) and protein (36g) resulted in faster wound healing, shortening of immobilisation period, and increased muscle strength in older malnourished hip fracture patients.^89^ A further study involving older hip fracture patients supplemented with CaHMB (2.1g) also observed improved muscle mass and function and prevention of the onset of sarcopenia.^90^ CaHMB plus amino acid supplementation in older adults infected with human immunodeficiency virus (HIV) reduced viral infection and muscle wasting.^91^ Findings from a recent umbrella review support the individual effects of the above RCTs by concluding there is high-quality evidence favouring supplementation of HMB (above the recommended daily intake) for increasing muscle mass in older adults.^92^ Therefore, the provision of CaHMB/vitamin D/protein in older adults with sarcopenia may be a viable option to enhance or preserve muscle mass and function.

Trace elements in the multi-nutrient supplement also contribute to improving immune function. Selenium supplementation (50, 100 or 200 µg/day Se-enriched yeast) of adults with suboptimal status is considered a safe adjuvant therapy against viral infections.^93^ In older adults, selenium supplement (100μg/day) improves the response to influenza vaccination and is accompanied by increased IFN-γ levels after vaccination.^94^ Zinc is important for the development and maintenance of cell-mediated immunity^95^ Zinc deficiency is associated with decreased immune responses with ageing, and improving zinc status can in part reverse immune dysfunction and reduce chronic inflammation associated with ageing.^96^ In addition, other nutrients, such as vitamins A, B, C and E, essential for T cell function, significantly improve the severity and mortality rate in COVID-19 patients.^97^

The main purpose of this study is to prepare aged-care residents against viral infections, improve their immune, muscle and respiratory function and quality of life. Therefore, this study may have significant health impacts that are broader than the preparation for or prevention of COVID-19. Outcomes from this study may provide evidence-based clinical care pathways to support scalable and pragmatic aged-care based nutrition support programs that reduce the severity of seasonal influenza or other viral infections.

### Conclusion

In conclusion, the Pomerium Study will determine the efficacy of a multi-nutrient supplement on immune, muscle and respiratory function and QoL of older adults in aged care. Further outcomes include a reduction in the incidence of COVID-19 and seasonal viral infections and their associated complications in supplemented participants. The study results may support the provision of multi-nutrient supplements to older adults in aged care prior to and during viral outbreaks as a strategy to reduce the onset and severity of viral infections.

## Data Availability

All data produced in the present study are available upon reasonable request to the authors

## DECLARATIONS

### Ethics approval and consent to participate

This study was approved by Melbourne Health Human Research Ethics Committee (Ref No. HREC/73985/MH-2021, ERM Ref No. RMH73985, Melbourne Health Site Ref No. 2021.115).

### Consent for publication

All the authors have consented to publish this protocol.

### Availability of data and material

Future datasets generated and/or analysed during the study will not be publicly available due privacy issues but will be available from the corresponding author on reasonable request.

### Competing interests

GD and SI serve as members of the Advisory Board of Abbott Australia. GD serves on the Scientific Advisory Board at TSI Pharmaceuticals. BK is currently supported by a research fellowship from TSI Pharmaceuticals. All the other authors declare that they have no competing interests.

### Funding

This study was funded by a grant from the 2020 Rare Cancers Rare Diseases and Unmet Need – COVID-19 grants program by the Australian Medical Research Future Fund (MRFF).

### Sponsor

Western Health (Melbourne, Australia)

### Role of study sponsor and funder

The sponsor and funder agency will not play any role in study design; collection, management, analysis, and interpretation of data; writing of the report; or the decision to submit the report for publication.

### Author contributions

GD conceived the trial and is the Chief Investigator in the MRFF grant. All authors participated in the design of the trial and read and approved the final manuscript.

## Notes

### Clinical Trial

ACTRN12621000420842

### Author Declarations

It is approved by Melbourne Health Human Research Ethics Committee (Ref No. HREC/73985/MH-2021, ERM Ref No. RMH73985, Melbourne Health Site Ref No. 2021.115)

## References

1. Leng SX, Margolick JB. Aging, sex, inflammation, frailty, and CMV and HIV infections. Cell Immunol 2020;348(1090-2163 (Electronic)):104024. doi: 10.1016/j.cellimm.2019.104024 [published Online First: 2019/12/18]

2. Cunha LL, Perazzio SF, Azzi J, et al. Remodeling of the Immune Response With Aging: Immunosenescence and Its Potential Impact on COVID-19 Immune Response. Front Immunol 2020;11(1664-3224 (Electronic)):1748. doi: 10.3389/fimmu.2020.01748 [published Online First: 2020/08/28]

3. Fulop T, Larbi A, Dupuis G, et al. Immunosenescence and Inflamm-Aging As Two Sides of the Same Coin: Friends or Foes? Front Immunol 2017;8(1664-3224 (Print)):1960. doi: 10.3389/fimmu.2017.01960 [published Online First: 2018/01/30]

4. Li H, Manwani B, Leng SX. Frailty, inflammation, and immunity. Aging Dis 2011;2(6):466–73. [published Online First: 2012/03/08]

5. Chen Y, Liu S, Leng SX. Chronic Low-grade Inflammatory Phenotype (CLIP) and Senescent Immune Dysregulation. Clin Ther 2019;41(3):400–09. doi: 10.1016/j.clinthera.2019.02.001 [published Online First: 2019/03/06]

6. Woods JL, Iuliano-Burns S, Walker KZ. Immunological and nutritional factors in elderly people in low-level care and their association with mortality. Immun Ageing 2013;10(1742-4933 (Print)):32. doi: 10.1186/1742-4933-10-32 [published Online First: 2013/08/07]

7. Sugg MM, Spaulding TJ, Lane SJ, et al. Mapping community-level determinants of COVID-19 transmission in nursing homes: A multi-scale approach. Sci Total Environ 2021;752(1879-1026 (Electronic)):141946. doi: 10.1016/j.scitotenv.2020.141946 [published Online First: 2020/09/06]

8. Iuliano S, Poon S, Wang XF, et al. Dairy food supplementation may reduce malnutrition risk in institutionalised elderly. British Journal of Nutrition 2017;117(1):142–47. doi: 10.1017/S000711451600461x

9. Nalin D. Immunosenescence and Severe Acute Respiratory Syndrome Coronavirus 2 Vaccine Development. The Journal of infectious diseases 2020;222(12):2114. doi: 10.1093/infdis/jiaa564 [published Online First: 2020/09/06]

10. Fagnoni FF, Vescovini R, Mazzola M, et al. Expansion of cytotoxic CD8+ CD28-T cells in healthy ageing people, including centenarians. Immunology 1996;88(4):501–7. doi: 10.1046/j.1365-2567.1996.d01-689.x [published Online First: 1996/08/01]

11. Pae M, Wu D. Nutritional modulation of age-related changes in the immune system and risk of infection. Nutr Res. 2017 May;41:14–35. doi: 10.1016/j.nutres.2017.02.001.

12. Adams RL, Broughton KS. Insulinotropic Effects of Whey: Mechanisms of Action, Recent Clinical Trials, and Clinical Applications. Ann Nutr Metab 2016;69(1):56–63. doi: 10.1159/000448665 [published Online First: 2016/08/17]

13. Vyas N, Kurian SJ, Bagchi D, et al. Vitamin D in Prevention and Treatment of COVID-19: Current Perspective and Future Prospects. J Am Coll Nutr 2021;40(7):632–45. doi: 10.1080/07315724.2020.1806758 [published Online First: 2020/09/02]

14. Liu BA, Gordon M, Labranche JM, et al. Seasonal prevalence of vitamin D deficiency in institutionalized older adults. Journal of the American Geriatrics Society 1997;45(5):598–603. doi: 10.1111/j.1532-5415.1997.tb03094.x [published Online First: 1997/05/01]

15. Arnljots R, Thorn J, Elm M, et al. Vitamin D deficiency was common among nursing home residents and associated with dementia: a cross sectional study of 545 Swedish nursing home residents. BMC geriatrics 2017;17(1):229. doi: 10.1186/s12877-017-0622-1 [published Online First: 2017/10/12]

16. Malnutrition and Effects of an Individualized Nutritional Intervention: An enable Study. Nutrients. 2021 Jun 24;13(7):2168. doi: 10.3390/nu13072168.

17. Deutz NE, Bauer JM, Barazzoni R, et al. Protein intake and exercise for optimal muscle function with aging: recommendations from the ESPEN Expert Group. Clinical nutrition (Edinburgh, Scotland) 2014;33(6):929–36. doi: 10.1016/j.clnu.2014.04.007 [published Online First: 2014/05/13]

18. Z Arnljots R, Thorn J, Elm M, Moore M, Sundvall PD. Vitamin D deficiency was common among nursing home residents and associated with dementia: a cross sectional study of 545 Swedish nursing home residents. BMC Geriatr. 2017;17(1):229. doi: 10.1186/s12877-017-0622-1.

19. Sim M, Zhu K, Lewis JR, et al. Association between vitamin D status and long-term falls-related hospitalization risk in older women. Journal of the American Geriatrics Society 2021(1532-5415 (Electronic)) doi: 10.1111/jgs.17442 [published Online First: 2021/09/11]

20. Hoong CWS, Huilin K, Cho S, et al. Are Adequate Vitamin D Levels Helpful in Fighting COVID-19? A Look at the Evidence. Horm Metab Res 2020;52(11):775–83. doi: 10.1055/a-1243-5462 [published Online First: 2020/09/18]

21. Radujkovic A, Hippchen T, Tiwari-Heckler S, et al. Vitamin D Deficiency and Outcome of COVID-19 Patients. 2020(2072-6643 (Electronic))

22. Cruz-Jentoft AJ, Sayer AA. Sarcopenia. Lancet (London, England) 2019;393(10191):2636–46. doi: 10.1016/S0140-6736(19)31138-9 [published Online First: 2019/06/07]

23. Cruz-Jentoft AJ, Bahat G, Bauer J, et al. Sarcopenia: revised European consensus on definition and diagnosis. Age and ageing 2019;48(1):16–31. doi: 10.1093/ageing/afy169 [published Online First: 2018/10/13]

24. Sepúlveda-Loyola W, Osadnik C, Phu S, Morita AA, Duque G, Probst VS. Diagnosis, prevalence, and clinical impact of sarcopenia in COPD: a systematic review and meta-analysis. J Cachexia Sarcopenia Muscle. 2020 Oct;11(5):1164–1176. doi: 10.1002/jcsm.12600.

25. Martínez-Arnau Fm, Buigues C, Fonfría-Vivas R, Cauli O. Respiratory Muscle Strengths and Their Association with Lean Mass and Handgrip Strengths in Older Institutionalized Individuals. J Clin Med. 2020 Aug 24;9(9):2727. doi: 10.3390/jcm9092727.

26. Okazaki T, Ebihara S, Mori T, Izumi S, Ebihara T. Association between sarcopenia and pneumonia in older people. Geriatr Gerontol Int. 2020 Jan;20(1):7–13. doi: 10.1111/ggi.13839.

27. Nishimura JM, Ansari AZ, D’Souza DM, et al. Computed Tomography-Assessed Skeletal Muscle Mass as a Predictor of Outcomes in Lung Cancer Surgery. Ann Thorac Surg 2019;108(5):1555–64. doi: 10.1016/j.athoracsur.2019.04.090 [published Online First: 2019/06/23]

28. Kim G, Kang SH, Kim MY, et al. Prognostic value of sarcopenia in patients with liver cirrhosis: A systematic review and meta-analysis. PloS one 2017;12(10):e0186990. doi: 10.1371/journal.pone.0186990 [published Online First: 2017/10/25]

29. Rovere-Querini P, Tresoldi C, Conte C, et al. Biobanking for COVID-19 research. Panminerva Med 2020 doi: 10.23736/S0031-0808.20.04168-3 [published Online First: 2020/10/20]

30. Ufuk F, Demirci M, Sagtas E, et al. The prognostic value of pneumonia severity score and pectoralis muscle Area on chest CT in adult COVID-19 patients. Eur J Radiol 2020;131:109271. doi: 10.1016/j.ejrad.2020.109271 [published Online First: 2020/09/18]

31. Kim JW, Yoon JS, Kim EJ, et al. Prognostic Implication of Baseline Sarcopenia for Length of Hospital Stay and Survival in Patients With Coronavirus Disease 2019. The journals of gerontology Series A, Biological sciences and medical sciences 2021;76(8):e110–e16. doi: 10.1093/gerona/glab085 [published Online First: 2021/03/30]

32. Feng Z, Zhao H, Kang W, et al. Association of Paraspinal Muscle Measurements on Chest Computed Tomography With Clinical Outcomes in Patients With Severe Coronavirus Disease 2019. The journals of gerontology Series A, Biological sciences and medical sciences 2021;76(3):e78–e84. doi: 10.1093/gerona/glaa317 [published Online First: 2020/12/24]

33. Kara O, Kara M, Akin ME, et al. Grip strength as a predictor of disease severity in hospitalized COVID-19 patients. Heart Lung 2021;50(6):743–47. doi: 10.1016/j.hrtlng.2021.06.005 [published Online First: 2021/07/05]

34. Cheval B, Sieber S, Maltagliati S, et al. Muscle strength is associated with COVID-19 hospitalization in adults 50 years of age or older. Journal of cachexia, sarcopenia and muscle 2021;12(5):1136–43. doi: 10.1002/jcsm.12738 [published Online First: 2021/08/08]

35. Welch C, Greig C, Masud T, et al. COVID-19 and Acute Sarcopenia. Aging Dis 2020;11(6):1345–51. doi: 10.14336/AD.2020.1014 [published Online First: 2020/12/04]

36. Piotrowicz K, Gasowski J, Michel JP, et al. Post-COVID-19 acute sarcopenia: physiopathology and management. Aging Clin Exp Res 2021;33(10):2887–98. doi: 10.1007/s40520-021-01942-8 [published Online First: 2021/07/31]

37. Murphy CH, Saddler NI, Devries MC, et al. Leucine supplementation enhances integrative myofibrillar protein synthesis in free-living older men consuming lower-and higher-protein diets: a parallel-group crossover study. Am J Clin Nutr 2016;104(6):1594–606. doi: 10.3945/ajcn.116.136424 [published Online First: 2016/12/10]

38. Moore DR, Churchward-Venne TA, Witard O, et al. Protein ingestion to stimulate myofibrillar protein synthesis requires greater relative protein intakes in healthy older versus younger men. The journals of gerontology Series A, Biological sciences and medical sciences 2015;70(1):57–62. doi: 10.1093/gerona/glu103 [published Online First: 2014/07/25]

39. Wilkinson DJ, Hossain T, Hill DS, et al. Effects of leucine and its metabolite beta-hydroxy-beta-methylbutyrate on human skeletal muscle protein metabolism. J Physiol 2013;591(11):2911-doi: 10.1113/jphysiol.2013.253203 [published Online First: 2013/04/05]

40. Oktaviana J, Zanker J, Vogrin S, et al. The Effect of beta-hydroxy-beta-methylbutyrate (HMB) on Sarcopenia and Functional Frailty in Older Persons: A Systematic Review. The journal of nutrition, health & aging 2019;23(2):145–50. doi: 10.1007/s12603-018-1153-y [published Online First: 2019/01/31]

41. Kirk B, Prokopidis K, Duque G. Nutrients to mitigate osteosarcopenia: the role of protein, vitamin D and calcium. Curr Opin Clin Nutr Metab Care 2021;24(1):25–32. doi: 10.1097/MCO.0000000000000711 [published Online First: 2020/11/06]

42. Kirk B, Feehan J, Lombardi G, et al. Muscle, Bone, and Fat Crosstalk: the Biological Role of Myokines, Osteokines, and Adipokines. Curr Osteoporos Rep 2020;18(4):388–400. doi: 10.1007/s11914-020-00599-y [published Online First: 2020/06/13]

43. Chanet A, Salles J, Guillet C, et al. Vitamin D supplementation restores the blunted muscle protein synthesis response in deficient old rats through an impact on ectopic fat deposition. J Nutr Biochem 2017;46(1873-4847 (Electronic)):30–38. doi: 10.1016/j.jnutbio.2017.02.024 [published Online First: 2017/04/27]

44. Verlaan S, Maier AB, Bauer JM, et al. Sufficient levels of 25-hydroxyvitamin D and protein intake required to increase muscle mass in sarcopenic older adults -The PROVIDE study. 2018(1532-1983 (Electronic))

45. Kositsawat J, Duque G, Kirk B. Nutrients with anabolic/anticatabolic, antioxidant, and anti-inflammatory properties: Targeting the biological mechanisms of aging to support musculoskeletal health. Experimental gerontology 2021;154(1873-6815 (Electronic)):111521. doi: 10.1016/j.exger.2021.111521 [published Online First: 2021/08/25]

46. Langkamp-Henken B, Wood SM, Herlinger-Garcia KA, et al. Nutritional formula improved immune profiles of seniors living in nursing homes. Journal of the American Geriatrics Society 2006;54(12):1861–70. doi: 10.1111/j.1532-5415.2006.00982.x [published Online First: 2007/01/03]

47. Akatsu H, Nagafuchi S, Kurihara R, et al. Enhanced vaccination effect against influenza by prebiotics in elderly patients receiving enteral nutrition. Geriatrics & gerontology international 2016;16(2):205–13. doi: 10.1111/ggi.12454 [published Online First: 2015/01/24]

48. Aspinall R, Lang PO. Interventions to restore appropriate immune function in the elderly. Immun Ageing 2018;15(1742-4933 (Print)):5. doi: 10.1186/s12979-017-0111-6 [published Online First: 2018/02/09]

49. Schulz KF, Altman Dg Fau - Moher D, Moher D. CONSORT 2010 statement: Updated guidelines for reporting parallel group randomised trials. 2010(0976-5018 (Electronic))

50. Iuliano S, Poon S, Robbins J, et al. Effect of dietary sources of calcium and protein on hip fractures and falls in older adults in residential care: cluster randomised controlled trial. BMJ (Clinical research ed) 2021;375:n2364. doi: 10.1136/bmj.n2364 [published Online First: 2021/10/22]

51. Gunawardene P, Bermeo S, Vidal C, et al. Association Between Circulating Osteogenic Progenitor Cells and Disability and Frailty in Older Persons: The Nepean Osteoporosis and Frailty Study. The journals of gerontology Series A, Biological sciences and medical sciences 2016;71(9):1124–30. doi: 10.1093/gerona/glv190 [published Online First: 2015/11/04]

52. Bhasin S, Travison TG, Manini TM, et al. Sarcopenia Definition: The Position Statements of the Sarcopenia Definition and Outcomes Consortium. Journal of the American Geriatrics Society 2020;68(7):1410–18. doi: 10.1111/jgs.16372 [published Online First: 2020/03/10]

53. Glendenning P, Inderjeeth CA. Vitamin D: Methods of 25 hydroxyvitamin D analysis, targeting at risk populations and selecting thresholds of treatment. Clinical Biochemistry 2012;45(12):901–06. doi: 10.1016/j.clinbiochem.2012.04.002

54. Phu S, Vogrin S, Al Saedi A, et al. Balance training using virtual reality improves balance and physical performance in older adults at high risk of falls. Clinical interventions in aging 2019;14:1567–77. doi: 10.2147/CIA.S220890 [published Online First: 2019/11/07]

55. Guralnik JM, Simonsick EM, Ferrucci L, et al. A short physical performance battery assessing lower extremity function: association with self-reported disability and prediction of mortality and nursing home admission. J Gerontol 1994;49(2):M85–94. doi: 10.1093/geronj/49.2.m85 [published Online First: 1994/03/01]

56. Phu S, Kirk B, Bani Hassan E, et al. The diagnostic value of the Short Physical Performance Battery for sarcopenia. BMC geriatrics 2020;20(1):242. doi: 10.1186/s12877-020-01642-4 [published Online First: 2020/07/15]

57. Miller MR, Hankinson J, Brusasco V, et al. Standardisation of spirometry. Eur Respir J 2005;26(2):319–38. doi: 10.1183/09031936.05.00034805 [published Online First: 2005/08/02]

58. Vellas B, Guigoz Y, Garry PJ, et al. The mini nutritional assessment (MNA) and its use in grading the nutritional state of elderly patients. Nutrition 1999;15(2):116–22. doi: Doi 10.1016/S0899-9007(98)00171-3

59. Abimanyi-Ochom J, Watts JJ, Borgstrom F, et al. Changes in quality of life associated with fragility fractures: Australian arm of the International Cost and Utility Related to Osteoporotic Fractures Study (AusICUROS). Osteoporosis international : a journal established as result of cooperation between the European Foundation for Osteoporosis and the National Osteoporosis Foundation of the USA 2015;26(6):1781–90. doi: 10.1007/s00198-015-3088-z [published Online First: 2015/03/21]

60. Beck AM, Christensen AG, Hansen BS, et al. Multidisciplinary nutritional support for undernutrition in nursing home and home-care: A cluster randomized controlled trial. Nutrition 2016;32(2):199–205. doi: 10.1016/j.nut.2015.08.009 [published Online First: 2015/11/11]

61. Sherwin AJ, Nowson CA, McPhee J, et al. Nutrient intake at meals in residential care facilites for the aged: validated visual estimation of plate waste. Australian journal of nutrition and dietetics 1998;55(4):188–93.

62. Schaffner W, van Buynder P, McNeil S, et al. Seasonal influenza immunisation: Strategies for older adults. International journal of clinical practice 2018;72(10):e13249. doi: 10.1111/ijcp.13249 [published Online First: 2018/09/15]

63. Doherty PC, Topham DJ, Tripp RA, et al. Effector CD4+ and CD8+ T-cell mechanisms in the control of respiratory virus infections. Immunol Rev 1997;159(0105-2896 (Print)):105–17. doi: 10.1111/j.1600-065x.1997.tb01010.x [published Online First: 1998/01/07]

64. Herrera MT, Gonzalez Y, Juarez E, et al. Humoral and cellular responses to a non-adjuvanted monovalent H1N1 pandemic influenza vaccine in hospital employees. BMC Infect Dis 2013;13(1471-2334 (Electronic)):544. doi: 10.1186/1471-2334-13-544 [published Online First: 2013/11/19]

65. Chapman TJ, Castrucci MR, Padrick RC, et al. Antigen-specific and non-specific CD4+ T cell recruitment and proliferation during influenza infection. Virology 2005;340(2):296–306. doi: 10.1016/j.virol.2005.06.023 [published Online First: 2005/08/02]

66. Schmidt T, Dirks J, Enders M, et al. CD4+ T-cell immunity after pandemic influenza vaccination cross-reacts with seasonal antigens and functionally differs from active influenza infection. Eur J Immunol 2012;42(7):1755–66. doi: 10.1002/eji.201242393 [published Online First: 2012/05/16]

67. Meydani SN, Leka LS, Fine BC, et al. Vitamin E and respiratory tract infections in elderly nursing home residents: a randomized controlled trial. JAMA 2004;292(7):828–36. doi: 10.1001/jama.292.7.828 [published Online First: 2004/08/19]

68. Girodon F, Lombard M, Galan P, et al. Effect of micronutrient supplementation on infection in institutionalized elderly subjects: a controlled trial. Ann Nutr Metab 1997;41(2):98–107. doi: 10.1159/000177984 [published Online First: 1997/01/01]

69. Kirk B, Mooney K, Vogrin S, et al. Leucine-enriched whey protein supplementation, resistance-based exercise, and cardiometabolic health in older adults: a randomized controlled trial. Journal of cachexia, sarcopenia and muscle 2021 doi: 10.1002/jcsm.12805 [published Online First: 2021/09/15]

70. Nguyen V, Mendelsohn A, Larrick JW. Interleukin-7 and Immunosenescence. J Immunol Res 2017;2017(2314-7156 (Electronic)):4807853. doi: 10.1155/2017/4807853 [published Online First: 2017/05/10]

71. Constantin D, Menon MK, Houchen-Wolloff L, et al. Skeletal muscle molecular responses to resistance training and dietary supplementation in COPD. Thorax 2013;68(7):625–33. doi: 10.1136/thoraxjnl-2012-202764 [published Online First: 2013/03/29]

72. Nutritional guidelines and menu checklist for residential and nursing homes. EFAD guidelines 2014

73. Best Practices for Nutrition, Food Service and Dining in Long Term Care Homes. Dietitians of Canada 2019

74. Caccialanza R, Laviano A, Lobascio F, et al. Early nutritional supplementation in non-critically ill patients hospitalized for the 2019 novel coronavirus disease (COVID-19): Rationale and feasibility of a shared pragmatic protocol. Nutrition 2020;74(1873-1244 (Electronic)):110835. doi: 10.1016/j.nut.2020.110835 [published Online First: 2020/04/14]

75. Alexander J, Tinkov A, Strand TA, et al. Early Nutritional Interventions with Zinc, Selenium and Vitamin D for Raising Anti-Viral Resistance Against Progressive COVID-19. Nutrients 2020;12(8) doi: 10.3390/nu12082358 [published Online First: 2020/08/14]

76. Panagiotou GA-O, Tee SA, Ihsan Y, et al. Low serum 25-hydroxyvitamin D (25[OH]D) levels in patients hospitalized with COVID-19 are associated with greater disease severity. 2020(1365-2265 (Electronic))

77. Goncalves-Mendes N, Talvas J, Duale C, et al. Impact of Vitamin D Supplementation on Influenza Vaccine Response and Immune Functions in Deficient Elderly Persons: A Randomized Placebo-Controlled Trial. Front Immunol 2019;10(1664-3224 (Electronic)):65. doi: 10.3389/fimmu.2019.00065 [published Online First: 2019/02/26]

78. Alavi Foumani A, Mehrdad M, Jafarinezhad A, et al. Impact of vitamin D on spirometry findings and quality of life in patients with chronic obstructive pulmonary disease: a randomized, double-blinded, placebo-controlled clinical trial. Int J Chron Obstruct Pulmon Dis 2019;14:1495–501. doi: 10.2147/COPD.S207400 [published Online First: 2019/07/31]

79. Rafiq R, Prins HJ, Boersma WG, et al. Effects of daily vitamin D supplementation on respiratory muscle strength and physical performance in vitamin D-deficient COPD patients: a pilot trial. Int J Chron Obstruct Pulmon Dis 2017;12:2583–92. doi: 10.2147/COPD.S132117 [published Online First: 2017/09/13]

80. Gold DR, Litonjua AA, Carey VJ, et al. Lung VITAL: Rationale, design, and baseline characteristics of an ancillary study evaluating the effects of vitamin D and/or marine omega-3 fatty acid supplements on acute exacerbations of chronic respiratory disease, asthma control, pneumonia and lung function in adults. Contemp Clin Trials 2016;47:185–95. doi: 10.1016/j.cct.2016.01.003 [published Online First: 2016/01/20]

81. Dedeyne L, Dupont J, Koppo K, et al. Exercise and Nutrition for Healthy AgeiNg (ENHANce) project -effects and mechanisms of action of combined anabolic interventions to improve physical functioning in sarcopenic older adults: study protocol of a triple blinded, randomized controlled trial. BMC geriatrics 2020;20(1):532. doi: 10.1186/s12877-020-01900-5 [published Online First: 2020/12/12]

82. Rondanelli M, Klersy C, Terracol G, et al. Whey protein, amino acids, and vitamin D supplementation with physical activity increases fat-free mass and strength, functionality, and quality of life and decreases inflammation in sarcopenic elderly. Am J Clin Nutr 2016;103(3):830–40. doi: 10.3945/ajcn.115.113357 [published Online First: 2016/02/13]

83. Rondanelli M, Cereda E, Klersy C, et al. Improving rehabilitation in sarcopenia: a randomized-controlled trial utilizing a muscle-targeted food for special medical purposes. Journal of cachexia, sarcopenia and muscle 2020;11(6):1535–47. doi: 10.1002/jcsm.12532 [published Online First: 2020/09/23]

84. Bauer JM, Verlaan S, Bautmans I, et al. Effects of a vitamin D and leucine-enriched whey protein nutritional supplement on measures of sarcopenia in older adults, the PROVIDE study: a randomized, double-blind, placebo-controlled trial. Journal of the American Medical Directors Association 2015;16(9):740–7. doi: 10.1016/j.jamda.2015.05.021 [published Online First: 2015/07/15]

85. Cramer JT, Cruz-Jentoft AJ, Landi F, et al. Impacts of High-Protein Oral Nutritional Supplements Among Malnourished Men and Women with Sarcopenia: A Multicenter, Randomized, Double-Blinded, Controlled Trial. Journal of the American Medical Directors Association 2016;17(11):1044–55. doi: 10.1016/j.jamda.2016.08.009 [published Online First: 2016/10/27]

86. Fan H, Hong B, Luo Y, et al. The effect of whey protein on viral infection and replication of SARS-CoV-2 and pangolin coronavirus in vitro. Signal Transduct Target Ther 2020;5(1):275. doi: 10.1038/s41392-020-00408-z [published Online First: 2020/11/26]

87. Hsieh LC, Chow CJ, Chang WC, et al. Effect of beta-hydroxy-beta-methylbutyrate on protein metabolism in bed-ridden elderly receiving tube feeding. Asia Pac J Clin Nutr 2010;19(2):200–8. [published Online First: 2010/05/13]

88. Wu H, Xia Y, Jiang J, et al. Effect of beta-hydroxy-beta-methylbutyrate supplementation on muscle loss in older adults: a systematic review and meta-analysis. Archives of gerontology and geriatrics 2015;61(2):168–75. doi: 10.1016/j.archger.2015.06.020 [published Online First: 2015/07/15]

89. Ekinci O, Yanik S, Terzioglu Bebitoglu B, et al. Effect of Calcium beta-Hydroxy-beta-Methylbutyrate (CaHMB), Vitamin D, and Protein Supplementation on Postoperative Immobilization in Malnourished Older Adult Patients With Hip Fracture: A Randomized Controlled Study. Nutr Clin Pract 2016;31(6):829–35. doi: 10.1177/0884533616629628 [published Online First: 2016/03/12]

90. Malafarina V, Uriz-Otano F, Malafarina C, et al. Effectiveness of nutritional supplementation on sarcopenia and recovery in hip fracture patients. A multi-centre randomized trial. Maturitas 2017;101(1873-4111 (Electronic)):42–50. doi: 10.1016/j.maturitas.2017.04.010 [published Online First: 2017/05/26]

91. Clark RH, Feleke G, Din M, et al. Nutritional treatment for acquired immunodeficiency virus-associated wasting using beta-hydroxy beta-methylbutyrate, glutamine, and arginine: a randomized, double-blind, placebo-controlled study. JPEN J Parenter Enteral Nutr 2000;24(3):133–9. doi: 10.1177/0148607100024003133 [published Online First: 2000/06/13]

92. Gielen E, Beckwee D, Delaere A, et al. Nutritional interventions to improve muscle mass, muscle strength, and physical performance in older people: an umbrella review of systematic reviews and meta-analyses. Nutr Rev 2021;79(2):121–47. doi: 10.1093/nutrit/nuaa011 [published Online First: 2020/06/03]

93. Goldson AJ, Fairweather-Tait SJ, Armah CN, et al. Effects of selenium supplementation on selenoprotein gene expression and response to influenza vaccine challenge: a randomised controlled trial. PloS one 2011;6(3):e14771. doi: 10.1371/journal.pone.0014771 [published Online First: 2011/03/30]

94. Ivory K, Prieto E, Spinks C, et al. Selenium supplementation has beneficial and detrimental effects on immunity to influenza vaccine in older adults. Clinical Nutrition 2017;36(2):407–15. doi: 10.1016/j.clnu.2015.12.003

95. Maares M, Haase H. Zinc and immunity: An essential interrelation. Arch Biochem Biophys 2016;611(1096-0384 (Electronic)):58–65. doi: 10.1016/j.abb.2016.03.022 [published Online First: 2016/03/30]

96. Wong CP, Magnusson KR, Sharpton TJ, et al. Effects of zinc status on age-related T cell dysfunction and chronic inflammation. Biometals 2021;34(2):291–301. doi: 10.1007/s10534-020-00279-5 [published Online First: 2021/01/05]

97. Beigmohammadi MT, Bitarafan S, Hoseindokht A, et al. Impact of vitamins A, B, C, D, and E supplementation on improvement and mortality rate in ICU patients with coronavirus-19: a structured summary of a study protocol for a randomized controlled trial. Trials 2020;21(1):614. doi: 10.1186/s13063-020-04547-0 [published Online First: 2020/07/08]

