## Appendix 1 for "Effects of 12 weeks of Multi-nutrient supplementation on the Immune and Musculoskeletal systems of Older Adults in Aged-Care (The Pomerium Study): Protocol for a Randomised Controlled Trial"

**Appendix 1.** Nutrition profile and composition of the multi-nutrient supplement (Source: Abbott Pharmaceuticals Australia)

| **NUTRIENT** | **UNIT** | Per 220 mL (one bottle) |
| --- | --- | --- |
| Energy | kcal | 330 |
|  | kJ | 1388 |
| Protein | g | 20 |
| Fat | g | 11 |
| Carbohydrate | g | 37 |
| Of which sugars | g | 15 |
| Fibre | g | 1.7 |
| Of which FOS | g | 1.7 |
| HMB | g | 1.2 |
| Water | g | 168 |
| Carnitine | mg | 40 |
| Choline | mg | 154 |
| **Vitamins** |  |  |
| Vitamin A (palmitate) | µg RE | 132 |
|  | IU | 440 |
| Vitamin A (beta carotene) | µg RE | 132 |
|  | IU | 1320 |
| Vitamin D3 | µg | 13 |
|  | IU | 500 |
| Vitamin E | Mg TE | 5.5 |
|  | IU | 8.1 |
| Vitamin C | M9 | 35 |
| Vitamin K1 | µg | 33 |
| Folic acid | µg | 77 |
| Vitamin B1 | mg | 0.57 |
| Vitamin B2 | mg | 0.70 |
| Vitamin B6 | mg | 0.66 |
| Vitamin B12 | µg | 1.4 |
| Niacin equivalent | mg | 6.6 |
| Pantothenic acid | mg | 2.4 |
| Biotin | µg | 13.2 |
| **Minerals** |  |  |
| Sodium | mg | 330 |
| Potassium | mg | 594 |
| Chloride | mg | 139 |
| Calcium | mg | 500 |
| Phosphorus | mg | 260 |
| Magnesium | mg | 55 |
| Iron | mg | 4.6 |
| Zinc | mg | 3.9 |
| Manganese | mg | 0.99 |
| Copper | µg | 539 |
| Iodine | µg | 48 |
| Selenium | µg | 20 |
| Chromium | µg | 19 |
| Molybdenum | µg | 33 |

**FAT INFORMATION**

**Fat distribution**

| **Fat** | **g/100 mL** |
| --- | --- |
| Saturated fatty acids | 0.45 |
| Monounsaturated fatty acids | 2.21 |
| Polyunsaturated fatty acids | 1.73 |
| Trans fatty acids | 0.00033 |
| Cholesterol | 0.0050 |

**Fat sources**

| **Source** | **Proportion of total fat (%)** |
| --- | --- |
| Corn oil | 34.70 |
| Canola oil | 61.60 |
| Lecithin | 3.70 |

**PROTEIN INFORMATION**

**Protein sources**

| **Source** | **Proportion of total protein (%)** |
| --- | --- |
| Whey protein concentrate (75% protein) | 10 |
| Isolated soy protein, identity preserved | 15 |
| Milk protein concentrate and/or isolate | 35 |
| Sodium caseinate – low viscosity | 40 |

Total kcal/nitrogen ratio: 103:1

Non-protein kcal/nitrogen ratio: 78:1

**CARBOHYDRATE INFORMATION**

**Carbohydrate profile**

| **Carbohydrate** | **g/100 mL** |
| --- | --- |
| Sugars (mono + disaccharides) | 7.00 |
| Monosaccharides | 0.30 |
| Disaccharides | 6.70 |
| Higher saccharides | 9.70 |
| Maltotriose | 1.50 |
| Fibre | 0.75 |

Typical lactose content: 2212 mg/L

**LIST OF INGREDIENTS IN DESCENDING ORDER**

Water, hydrolysed corn starch, sucrose, VEGETABLE OIL (canola oil, corn oil), sodium caseinate, milk protein, soy protein isolate, whey protein concentrate, MINERALS (potassium citrate, sodium citrate, calcium phosphate tribasic, magnesium carbonate, potassium chloride, ferrous sulphate, zinc sulphate, manganese sulphate, cupric sulphate, sodium molybdate, potassium iodide, chromium chloride, sodium selenate), fructo-oligosaccharides, CaHMB (calcium β-hydroxy-β-methylbutyrate monohydrate), flavouring, soy lecithin, cellulose, choline chloride, VITAMINS (ascorbic acid, dl- alpha tocopheryl acetate, niacinamide, calcium pantothenate, beta carotene, pyridoxine hydrochloride, thiamine hydrochloride, riboflavin, vitamin A palmitate, folic acid, phylloquinone, vitamin D_3_, biotin, cyanocobalamin), L-carnitine, sodium carboxymethyl cellulose, and gellan gum.

May contain potassium phosphate dibasic and sodium chloride.
