## Appendix 2 for "Effects of 12 weeks of Multi-nutrient supplementation on the Immune and Musculoskeletal systems of Older Adults in Aged-Care (The Pomerium Study): Protocol for a Randomised Controlled Trial"

**Appendix 2,** Human Cytokine/Chemokine Milliplex Panel include the following analytes:

- sCD40L
- EGF
- FGF-2
- Flt-3 ligand
- Fractalkine
- G-CSF
- GM-CSF
- GRO IL-4
- IL-5
- IL-6
- IL-7
- IL-8
- IL-9
- IL-10
- IL-12 (p40)
- IL-12 (p70)
- IL-13
- IL-15
- IL-17A
- IP-10
- MCP-1
- MCP-3
- MDC (CCL22)
- MIP-1α
- MIP-1β
- PDGF-AB/BB
- RANTES
- TGF-α
- TNF-α
- TNF-β
- VEGF
- Eotaxin/CCL11
- PDGF-AA
- IFN-α2
- IFN-γ
- IL-1α
- IL-1β
- IL-1ra
- IL-2
- IL-3
